# Framingham risk score prediction at 12 months in the STANDFIRM randomised control trial

**DOI:** 10.1101/2023.09.05.23295104

**Authors:** Thanh G Phan, Velandai K Srikanth, Dominique A Cadilhac, Mark Nelson, Joosup Kim, Muideen T Olaiya, Sharyn M Fitzgerald, Christopher Bladin, Richard Gerraty, Henry Ma, Amanda G Thrift, the Shared Team Approach between Nurses and Doctors For Improved Risk factor Management (STANDFIRM) Collaborators

## Abstract

**Background:** The Shared Team Approach between Nurses and Doctors For Improved Risk factor Management (STANDFIRM, ACTRN12608000166370) trial was designed to test the effectiveness of chronic disease care management for modifying the Framingham risk score (FRS) among patients with stroke or transient ischemic attack. The primary outcome of change in FRS between baseline and 12 months was not met. We aimed to determine characteristics of participants at baseline that predict reduction in FRS at 12 months and whether future FRS is predetermined at the time of randomization

**Method:** Data included 35 variables encompassing demographics, risk factors, psychological, social and education status, and laboratory tests. Five supervised machine learning (ML) methods were used: random forest (RF), extreme gradient boosting (XGBoost), support vector regression (SVR), multilayer perceptron artificial neural network (MLP) and K-nearest neighbor (KNN). We split data for training (80%, n=406) and testing (20%, n=102).

**Results:** Training and test data were evenly matched for age, sex, baseline and 12-month FRS. Following tuning of the five ML methods, the optimal model for predicting FRS at 12 months was SVR (R^2^=0.763, root mean squared error or RMSE=8.52). The five most important variables for SVR were: baseline FRS, age, male sex, sodium/potassium excretion and proteinuria. All ML methods were poor at determining change in FRS at 12 months (R^2^<0.161).

**Conclusion:** Our findings suggest that change in FRS as an endpoint in trials may have limited value as it is largely determined at baseline. In this cohort, Support Vector Regression was the optimal method to predict future but not change in FRS.

## Introduction

Stroke is the second leading cause of death and third leading cause of disability worldwide and results in significant economic and societal cost^1^. In Australia, the lifetime risk of stroke from the age of 25 years was one in five ^2^. Despite trials providing evidence for the effectiveness of antithrombotic, antihypertensive medication and statins for stroke prevention, a pathway for consistently implementing these findings, as well as individualising therapy, is lacking^3–5^. In Australia, a Medicare item known as chronic disease care management plan was implemented to aid general practitioners to manage patients with chronic disease, including stroke. The Shared Team Approach between Nurses and Doctors For Improved Risk Factor Management (STANDFIRM) trial was designed to test the use of chronic disease care management plan to modify risk factors among patients with ischemic and hemorrhagic stroke (trial registration ACTRN12608000166370)^6^. The intervention arm included the use of chronic disease care plans generated by a stroke physician and supported by a nurse educator working in collaboration with patients and their family doctor.

For the primary outcome of this trial, there was no difference in the Framingham risk score (FRS 0.04, 95% CI −1.7 – 1.8) between the randomized groups at 12 months after randomization. This outcome might have occurred because of the high frequency of secondary stroke prevention medications prescribed at hospital discharge^6^. Several similar trials in patients with stroke have also obtained negative results when using change in FRS as outcome ^7–10^. The reasons underlying this finding remain unclear, but the baseline variables (at the time of recruitment in hospital or stroke clinic) in our cohort could potentially determine future FRS and if that is the case then it has implications for using the FRS in clinical trials of secondary stroke prevention.

In this exploratory (post-hoc) analysis, and applying a range of machine learning methods, we used baseline variables to predict FRS at 12 months and change in the score from baseline. This approach was performed to test the hypothesis that future FRS is predetermined at baseline or time of randomization. Furthermore, we wanted to evaluate the role of the other covariates in the outcome of FRS. Initially, we had considered using linear regression analysis as clinicians are comfortable with the outputs that are expressed in terms of regression coefficients and p-values^11^. A potential drawback in regression analyses is that only a small number of covariates collected during clinical trials are used and a large portion of the collected data are often not used. This issue is related to the underlying architecture of the linear regression method versus the non-linear supervised machine learning methods used here (see Supplementary Material for further details)^12^. The draw-back to using machine learning tools is their poor standing as ‘black box’ and, by inference, lack of model interpretability^12^. Recent addition of explainable machine learning tools, based on cooperative game theory, permit unravelling of the ‘mystery’ of machine learning. This implementation seeks to explain the model in a way analogous to distributing the profit resulting from coalition of workers relative to their individual contribution ^13, 14^.

## Method

### Data

The methodology, primary outcome, and secondary analyses of this trial have been published^6, 15–17^. There were 35 covariates (features) available for analyses including demographic, social, anxiety, depression and education status (9 variables), dietary and alcohol intake (8 variables), stroke risk factors (9 variables), blood and urine electrolytes (8 variables) and FRS. The FRS was calculated using the method by D’Agostino with higher score indicating a greater risk of death from cardiovascular disease^18^. Change in FRS was defined as difference between scores at baseline and 12 months.

### Statistical Analysis

All analyses were based on scikit-learn library in Python programming language version 3.7.3 ^19^. In this section, we provide a very brief overview of Support Vector Regression (SVR) while tree-based methods (extreme gradient boost machine/XGBoost and random forest/RF), multi-later perceptron (MLP) neural network and K-Nearest Neighbour regression are covered in the Supplementary Material ^12, 19^. SVR is a machine learning method which uses of kernel manipulation and support vectors (around margin of the plane to reduce errors to an acceptable limit) to perform regression. This idea can be interpreted as the hyperplane separating two classes of data in two-dimensional space is a line. By contrast, a plane is better suited to describe transformed data with three or more dimensions. The idea in SVR is that data is easier to separate once transformed (kernel manipulation) into higher dimension space compared to its original low dimension native space. The fitted line arising from the margin of the data is analogous to fitting a regression line based on minimising least squares^19^. However, SVR differs from linear least squares regression in that it tries to minimise the errors to within an acceptable range.

An issue with using a large number of covariates for a given dataset is multicollinearity or relatedness of some of the covariates. We evaluate effect of multicollinearity on the outcome in several ways. Firstly, we removed highly correlated variables from the correlation matrix (Supplementary Figure 1) and calculated the variance inflation factors. The correlation matrix of the covariates can provide an indication of the presence of multicollinearity (Supplementary Figure 1)^12^. The sensitivity analysis is performed by first reducing variables with high (0.7) and then by those with moderate correlation (0.5) (Supplementary Figure 2).

The pre-processing steps included removal of outliers based on the concept of neighbourhood i.e. keep data points which are adjacent (neighbours) and remove data points which are far from the rest of the group defined by distance. This was followed by scaling of the data to ensure that all the data had the same range. Next, the scaled data were randomly split into training (80%) and test sets (20%). This is a standard approach in machine learning as it enables the model that we develop to be tested in a dataset that has not been used in development of the model. Each method was put through a tuning process including k-fold cross validation of the training data to search for the optimal parameters to run in the analyses (for the steps in the process see the Supplementary Material). The optimal model was chosen based on the minimal sum of squared errors for each method. Variable importance was determined by SHAP (SHapley Additive exPlanation) values using the SHAP library ^14^. The Shapley value is a dimensionless average measure of the marginal gain calculated for each of the covariate from all possible permutations of covariates with respect to the model without that covariate ^13^. The SHAP values produced here are additive combination of Shapley values and other local interpretable machine methods and hence we have used the term SHAP values in the paper^14^. To enable understanding of the covariates, dependence plots of interactions between covariates and each other are provided. Finally, the covariates identified as important from machine learning can be considered as the first step in feature extraction for linear regression.

## Results

The demographics of the STANDFIRM cohort have been published previously^6^. The training group (n=406) and test group (n=102) data were evenly matched in terms of baseline age (68.63±13.04 vs 66.56±13.89), sex, mean systolic blood pressure, creatinine, HDL cholesterol, body mass index, FRS and 12-month FRS (Table 1). There was a trend to difference in the sodium to potassium excretion between the 2 groups (p=0.07). The correlation matrix among the 35 covariates shows that variables having high correlation with baseline FRS were age (0.53), systolic blood pressure (0.52), male sex (0.42; Supplementary Figure 1). Age was positively correlated with systolic blood pressure (0.37) and negatively with creatinine excretion (−0.38). Male sex was correlated with creatinine excretion (0.49), body weight (0.28), alcohol (0.25) and salt intake (0.22). For the sensitivity analysis, we have reduced the number of covariates down to 32 when using a correlation coefficient <0.70 and 23 when using a correlation coefficient <0.50 (Supplementary Figure 2)

**Table 1:** Patient characteristics of the training and test datasets.

| Covariate | Train<br>n=406 | Test<br>n=102 | P value |
| --- | --- | --- | --- |
| FRS baseline score [mean ± SD] | 23.54± 15.01 | 22.83± 17.02 | 0.67 |
| FRS 12 months score [mean ± SD] | 24.67±15.59 | 23.68±17.58 | 0.56 |
| Age, years [mean ± SD] | 68.63±13.04 | 66.56±13.89 | 0.16 |
| Male, % | 0.67 | 0.63 | 0.45 |
| Ratio of sodium to potassium excretion, ratio [mean ± SD] | 2.11±0.95 | 2.31±0.94 | 0.07 |
| Serum Glucose, mmol/l [mean ± SD] | 5.57± 1.90 | 5.47 ±1.47 | 0.63 |
| Creatinine Excretion, micromol/L [mean ± SD] | 10.39± 3.79 | 10.34± 4.03 | 0.91 |
| Proteinuria, g/L, [mean ± SD] | 0.13± 0.34 | 0.14± 0.35 | 0.86 |
| Fruit consumption, per day [mean ± SD] | 1.61±1.13 | 1.42±1.03 | 0.12 |
| Systolic Blood Pressure, mmHg [mean ± SD] | 136.70±20.39 | 135.29±20.29 | 0.53 |

FRS, Framingham risk score; SD, standard deviation

### Choosing Machine learning model

Following tuning, the optimal model was SVR: SVR (R^2^=0.763, RMSE or root mean squared error=8.52), XGBoost (R^2^=0.690, RMSE=9.74), MLP (R^2^=0.707, RMSE=9.46), RF (R^2^=0.671, RMSE=10.03) and KNN (R^2^=0.464, RMSE=12.81). Plots of ground truth versus FRS at 12 months show that the observed (actual) and predicted values tended to vary with each other for SVR (Figure 1a). As such SVR was the better method for this data. By contrast, the predicted values for 12-month FRS were much lower than the observed values for XGBoost, RF and KNN (Figure 1). The time taken to tune the models was longest for MLP (577.4 minutes) and shortest for SVR (14.4 sec). These parameters indicate that SVR is the optimal model for this dataset followed by MLP and XGBoost.

**Figure 1:**
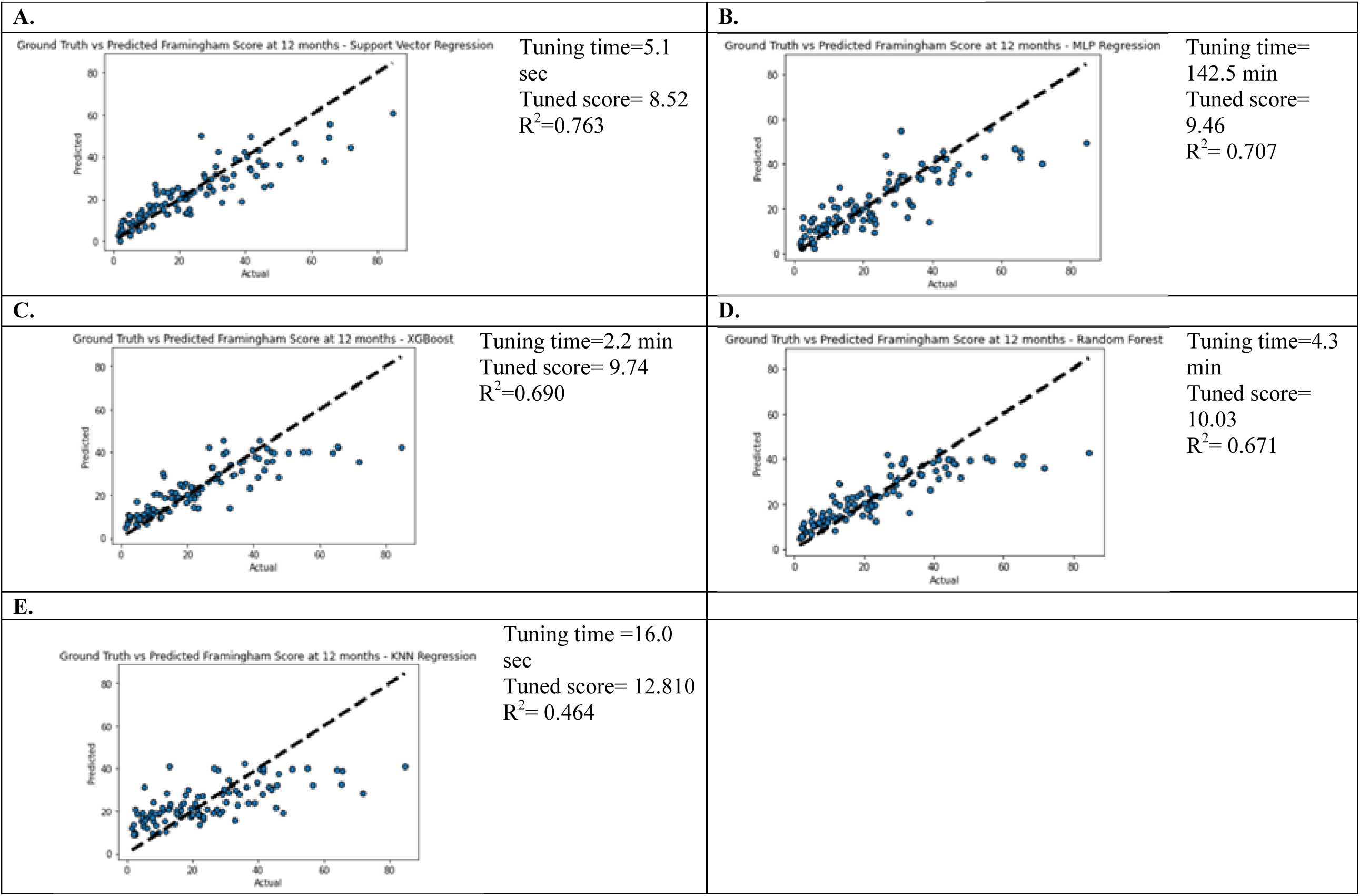
Model fitting: Plot of ground truth versus machine learning predicted values. The tuning time to optimise each machine learning method is shown on the panel. It was longest for multilayer perceptron (MLP) and shortest for support vector regression (SVR). There was deviation of the predicted from observed FRS values with random forest (RF), extreme gradient boost (XGBoost) and K-nearest neighbor (KNN) compared to SVR and MLP. The tuning time to optimise each machine learning method is shown on the panel. It was longest for multilayer perceptron (MLP) and shortest for support vector regression (SVR). There was deviation of the predicted from observed FRS values with random forest (RF), extreme gradient boost (XGBoost) and K-nearest neighbor (KNN) compared to SVR and MLP.

The five most important variables defined by dimensionless SHAP values for SVR were: baseline FRS (SHAP=8.84, 41.44%), age (SHAP=1.98, 9.28%), male (SHAP=1.87, 8.76%), ratio of urinary sodium to potassium excretion (SHAP=0.93, 4.36%), creatinine excretion (SHAP=0.61, 2.85%) (Figure 2). Figure 2 is the summary plot for SHAP values for all covariates and provides global interpretability. The summary plot (Figure 2a) showed that high baseline FRS, older age, or male sex are associated with high 12-month FRS. The sensitivity analyses for the different thresholds of correlation coefficient are provided in Figure 3. In essence, the R^2^ drop down to 0.728 at a correlation coefficient threshold of 0.50. Plots for the other machine learning methods illustrate the different extent to which the variables are used by each method (Supplementary Figure 3).

**Figure 2:**
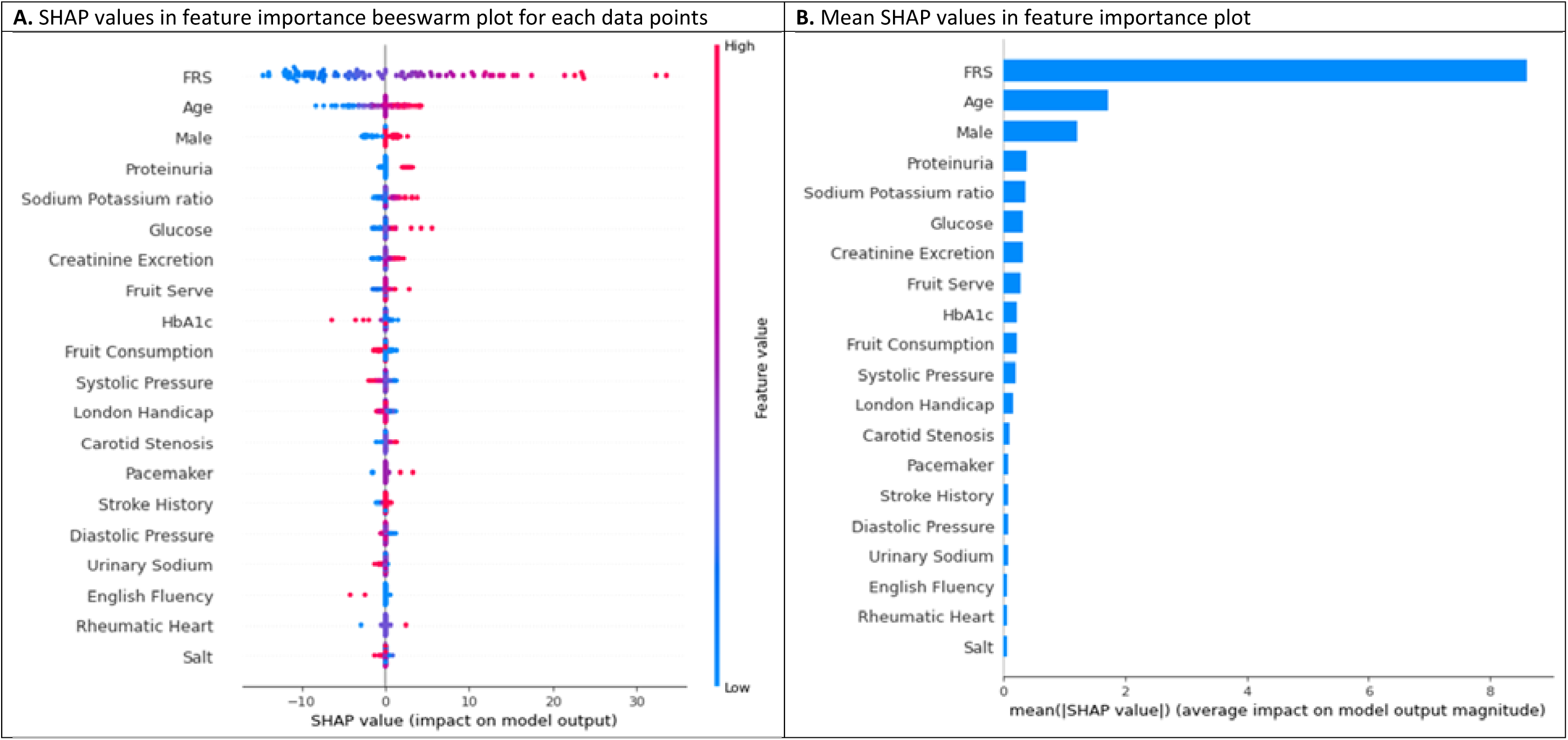
Global interpretability: SHAP values and covariates for support vector regression (SVR) The SHAP plot show that the covariates with the largest SHAP values are non-modifiable. FRS shift the prediction by 8.77 points, age by 2.17 points, male by 1.78 points, Sodium potassium ratio by 0.91 points. The dimensionless SHAP value is on the X-axis and the Y-axis represent the covariates ordered from highest to lowest importance. The color appears as a range for continuous scale and as 2 colors for binary values. FRS, Framingham risk score; IHD, ischemic heart disease, AF, atrial fibrillation; HbA1c, glycosylated hemoglobin.

**Figure 3:**
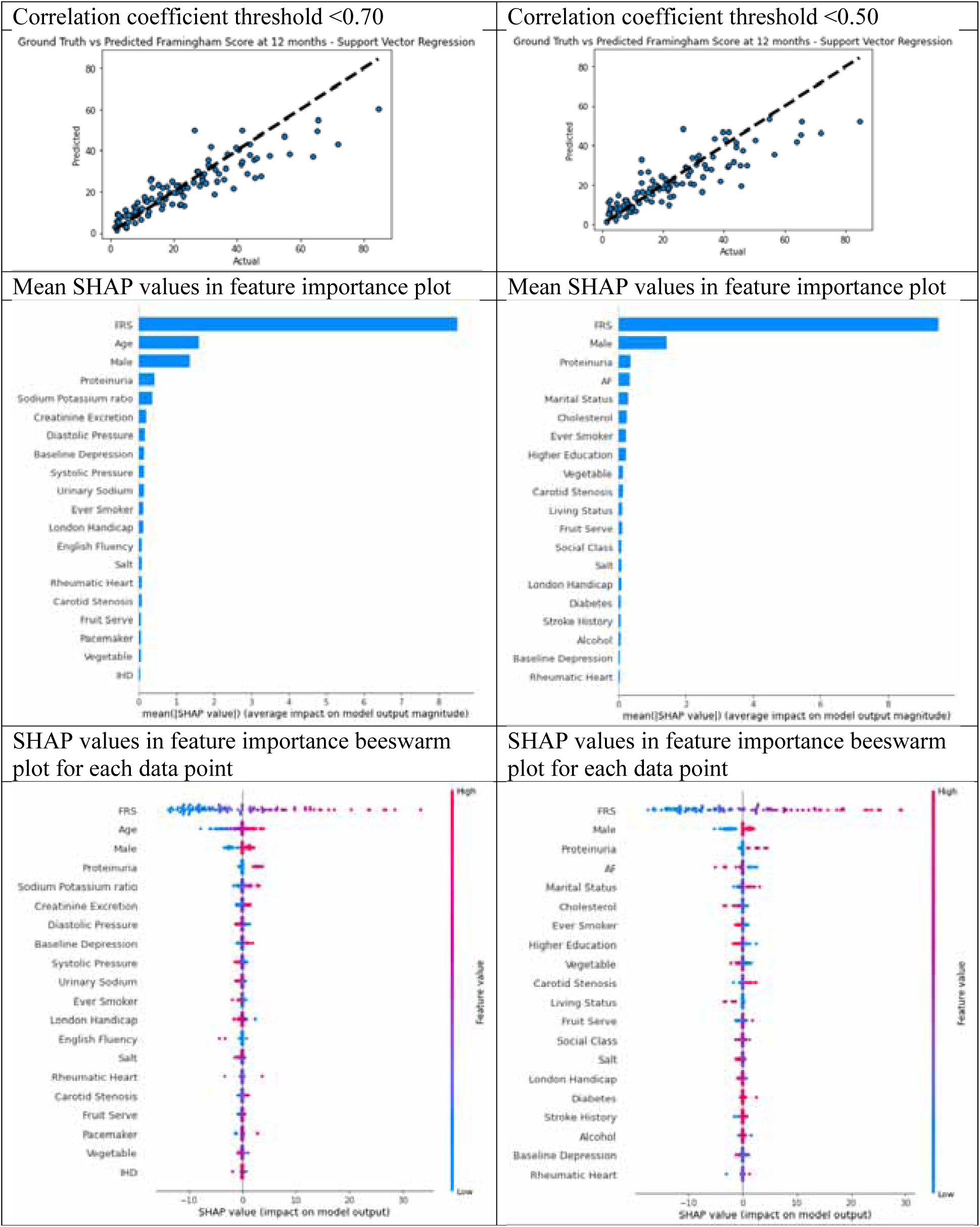
Global interpretability: Sensitivity analysis at correlation coefficient threshold of 0.7 and 0.5 The R^2^ drops from 0.770 for 32 variables data with down to 0.728 with 23 variables. FRS, Framingham risk score; IHD, ischemic heart disease, AF, atrial fibrillation; HbA1c, glycosylated hemoglobin.

### Dependence plot – interaction effect

The dependence plots for the 5 variables with the highest SHAP values are shown in Supplementary Figure 4. It shows interaction between FRS with alcohol, age with urinary potassium, male with systolic blood pressure, sodium potassium ratio with fruit consumption and proteinuria with FRS.

### Local interpretability

Local interpretability is provided in Figure 4 with the waterfall plot where covariates with the largest SHAP values affecting an individual’s prediction at the top and the covariates with the least prediction at the bottom. This style of plot is provided for the cases in which the difference between predicted and observed FRS is greater than 19 points (extreme points on histogram analysis, data not shown). The insight here is that FRS plays a major role in shifting the expected value to the final prediction for the individual patient on display in the Figure 4. while the next most important covariates rotate between age, male, proteinuria. The next level alternates among male, age, hypercholesterolemia and sodium potassium ratio. Five of these 6 patients were females (indicated by the negative value).

**Figure 4:**
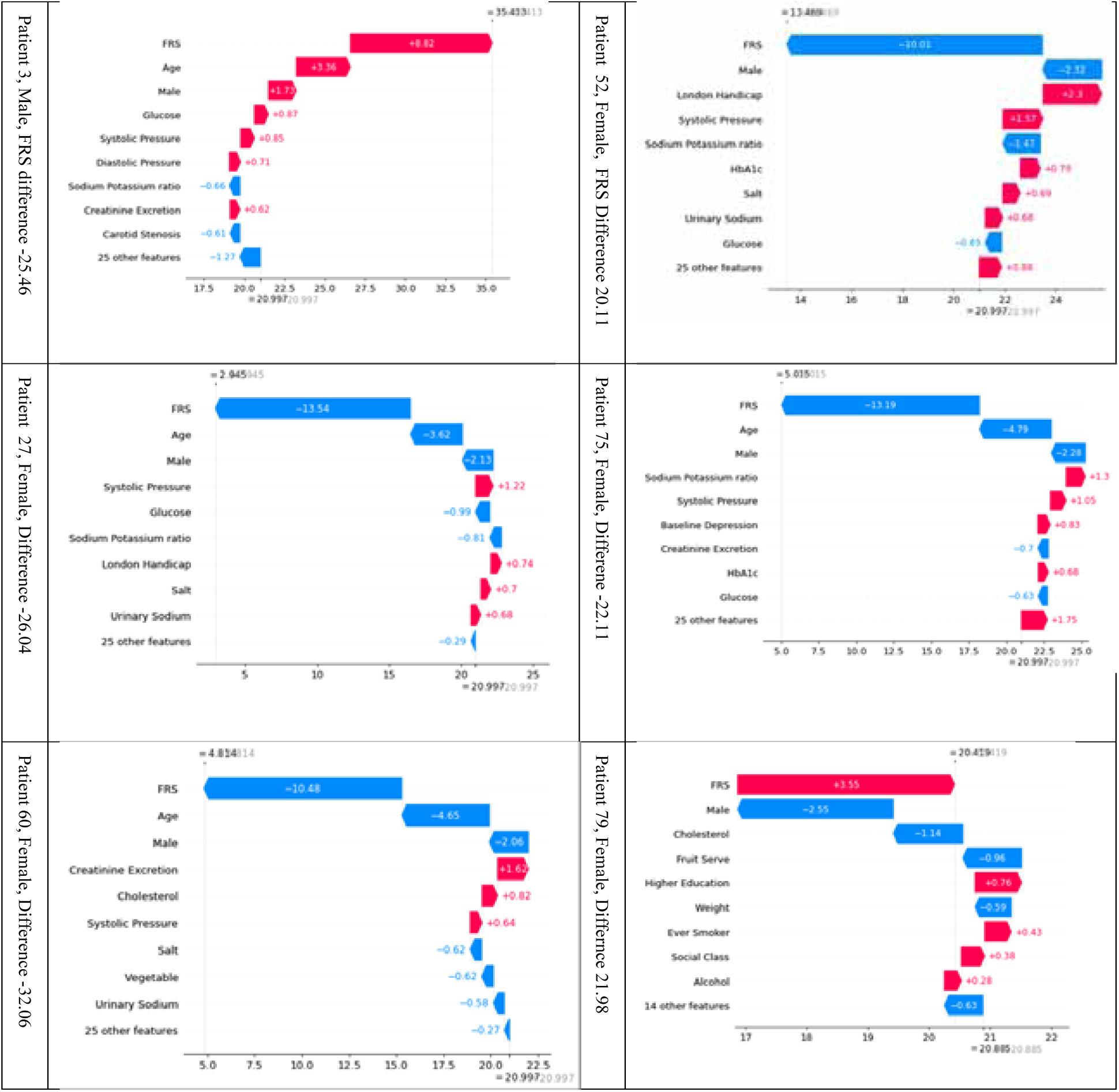
Local interpretability: Waterfall plot showing individual patient prediction in Test data illustrates the prediction for individual patients with greater than 19-point difference in predicted and observed FRS values. The expected values from the model for each patient is provided below the x-axis and the observed value is at the top. The plot show how the covariates move towards the prediction for that patient based on the SHAP values of each covariate, A feature of this waterfall plot is that the covariates with the largest contribution is listed at the top and the ones with the least contribution are listed at the bottom. The direction of the shift is provided in the Figure. Red color implies the shift is in the positive direction and blue color for shift in the negative direction.

### Linear regression

The five variables with the largest SHAP value from SVR were used in linear regression. The findings were: FRS (β=0.576, 95% CI 0.490-0.662, p <0.001), age (β =0.266, 95% CI 0.177-0.354, p <0.001), male (β=5.690, 95% CI 3.384-7.998, p <0.001), ratio of sodium to potassium excretion (β=1.214, 95% CI 0.227-2.200, p=0.016) and proteinuria (β=3.073, 95% CI 0.254 −5.892, p=0.03). The model R^2^ was 0.631.

### Change in FRS

In the trial cohort, all machine learning models performed poorly at predicting change in the FRS, with all R^2^ <0.161 (Supplementary Figure 5). Consequently, the SHAP values of the covariates are not displayed.

### Removal of baseline FRS from model

When the baseline FRS is removed from the data, the R^2^ for SVR dropped to 0.523 but the predicted values underestimate the higher observed values (Supplementary Figure 6). The top 5 features for SVR are older age, male, hypercholesterolemia, higher education, and ever-smoking.

## Discussion

In this analysis, we used various machine learning approaches to evaluate the large data available at the start of the patient journey to determine how well the 12-month FRS can be predicted. This is an issue critical to the planning of future trials based on using the FRS as an endpoint. The machine learning methods could predict future FRS with accuracy but not predict change in the FRS score. Importantly, three of the five most important variables associated with 12-month FRS were not modifiable. Our findings that the FRS outcome at 12 months is largely predetermined has large implications for conducting clinical trials undertaken to modify the risk of stroke as a main endpoint in trials. Consequently, FRS or change in FRS may not be an optimal choice as an endpoint.

### STANDFIRM trial

Practice appears to have changed considerably between planning and completion of the STANDFIRM trial. The trial was predicated on poor uptake of secondary stroke prevention in the community^20, 21^ and that addressing the risk factors through a structured program would reduce the 10-year cardiovascular risk ^15^. In one of these prior studies conducted in general practice, only 42% of patients with stroke or TIA were on secondary prevention therapy such as antithrombotic, antihypertensive medications and statins^21^, with only 27% of these patients being rated as being at high risk of stroke or TIA by their general practitioners ^21^.

Investigators from the North East Melbourne Stroke Incidence Study (NEMESIS) reported greater use of antihypertensive drugs at 10 years among those commencing these medications at hospital discharge (256 patients followed up at 10 years from an original cohort of 1,242 patients)^22^ This supported the notion that long-term adherence would be better in those commencing preventative medications at the time of hospital discharge. Observations from the STANDFIRM trial that >80% of patients were on secondary prevention therapy at the time of recruitment^6^, provides evidence for a significant change in clinical practice between the NEMESIS in 1997-99 and commencement of the STANDFIRM trial in 2013^6^. In accordance with this, selected centres in Australia and around the world have well structure outpatient clinics including TIA clinics and low rate of stroke recurrence^23^.

### Change in Framingham Risk Score

Patients were included in this study if they had a diagnosis of stroke and thus the FRS^18^ and its change were used to identify the effect of secondary prevention. With that in mind, the caveat to the findings from this analysis is that the FRS was used in a secondary prevention setting among hospitals classified as tertiary referral and do not necessarily invalidate the FRS approach in a specific setting of primary care and or for primary prevention of cardiovascular disease^18^. We were unable to develop models to predict change in FRS between baseline and 12 months. This finding is consistent with our observation that non-modifiable contributes to most of the total explainable FRS at 12 months (Figure 2). Serum creatinine, its excretion and proteinuria reflect renal function and have been identified as a risk factors for cardiovascular outcome and stroke recurrence^24, 25^. However, it’s not clear if these markers of renal function are reversible^26^. Sodium potassium ratio has been identified as risk factor for hypertension and potassium has physiologic role in decreasing sodium reabsorption. A recent metaanalysis of randomised control trials found a non-linear relationship between potassium supplement intake and systolic or diastolic blood pressure^27^. These investigators identified selected group in which excessive potassium intake can be detrimental. Glycemic control, systolic blood pressure and fruit consumption are potentially modifiable but individually they make up a small percentage of the SHAP values. This SHAP analysis suggest that if risk factor modification is to work, it should simultaneously address these multiple variables. Variables such as social demographics and level of education could be considered to be non-contributory based on the analysis of this cohort.

### Explaining machine learning

Machine learning is considered a black box and hence is less often used than classical regression methods. We have tried to circumvent this approach by using an interpretable machine learning (library SHAP) tool to aid interpretation of the model^14^. This method is agnostic to the method in question and can be applied to regression or classification analyses^35^. A caveat is that the explanation by SHAP is still dependent on us choosing the optimal machine learning method. As such we have created a ‘tournament’ style evaluation of the different machine learning method for this data. In this study we have provided global interpretability (average SHAP values in Figures 2, Supplementary Figure 3 and bees warm plot of individual data point in Figure 3) to understand how the model work. The local interpretability (waterfall plots in Figure 4) illustrates how the machine learning model is applied to predict outcome for the individual patient.

### Implication for use of FRS in clinical trials

Investigators of a Canadian trial found significant changes in FRS at 1 year for patients in the primary prevention arm but not those in the secondary prevention arm (n=296)^8^. By contrast, Japanese investigators did not observe a change in FRS in their trial of disease management plan (n=321)^7^. These stroke trials have been performed with relatively small sample size compared to cardiac trials ^27^. Secondary prevention trials for coronary artery disease, targeting lifestyle modification in the form of smoking cessation, increase physical activity and alcohol reduction, have shown a reduction in all-cause mortality ^27^. These findings potentially suggest that designing trials targeting lifestyle modification in stroke should replicate the cardiovascular trials with large sample size and with clinical endpoints.

Outside the setting of trials targeting change in FRS, other trials have incorporated a variety of methods to target post stroke blood pressure control in the community; these have been negative^9, 10, 28–30^. The setting of one of these studies was similar to the STANDFIRM trial, in which low-income patients in Southern California received a chronic care plan in a team setting, regardless of insurance status ^28^. This trial had no difference in blood pressure between the two groups at the end of the trial, but the intervention group had a greater self-reported reduction of salt intake and greater reduction in measured C-reactive protein than the usual care group^28^. In contrast, selected trials targeted at clinical endpoints (combined cardiovascular endpoints) rather than the FRS or medication compliance have been successful ^31^. Examples include the Austrian STROKE-CARD trial which comprised an e-tool and multidisciplinary team visit at 3 months to address risk factor modification.

### Clinical trial data

Similar to the STANDFIRM trial, often only a subset of the data collected in a clinical trial is used in regression analysis resulting in limited exploration of all the available data^12, 32^. This issue is likely related to the potential for overfitting regression analyses when a large number of covariates are used ^11^. Such issues are not always resolved by stepwise regression strategy^32^. In regression analyses, ‘rules’ have been proposed around the trade-off between number of covariates to sample size ^33^. Therefore, the possible contribution of variables such as anxiety, depression, level of education, fluency in English and other data regarding urinary electrolytes have not been incorporated into one global test. Because of the negative primary outcome of STANDFIRM, we have undertaken this post-hoc analysis to determine whether future trials should be conducted using the FRS as an outcome and the role of the other variables ^6^. To enable use of all the information available at the onset of the trial a different approach to regression analysis was required to determine future FRS with one solution being to use machine learning. In the medical literature machine learning methods are increasingly used as tool for classification^34^. These tools can also be used to perform regression tasks when the outcome is a continuous variable^12^.

### Structured approach to analysis

Among the machine learning methods available for tabular (structured) data, XGBoost is often seen as the go-to machine learning tool as it has been used successfully in competitions^19^. In this analysis, we carefully tuned different machine learning models and undertook a comparison between these methods to determine the best tool for the given data^19^. For this dataset, the finding that SVR was the best tool following tuning is critical as one should not assume that there is one general go-to machine learning tool for all data^12, 19^. MLP is an example of neural network with deep learning architecture in having multiple layers but it should not always be assumed to be better than other methods for tabular data such as the one here. In this cohort, MLP was computationally expensive and took a considerably longer time than the other methods^28^.

Multicollinearity or relatedness affects the accuracy of the coefficients of linear regression and the associated standard errors^12^. The authoritative book on machine learning methods cautioned against placing excessive trust on the regularization parameter in modulating multicollinearity as the regularization parameter is directed at all observations and not on the subspace in building of the hyperplane ^12^. In this analysis, removal of correlated terms did not improve the performance of tree-based method for the STANDFIRM data. MLP uses non-linear projection method to create multiple weights which are passed through the layers of neural network. In this way, neural network such as MLP are said to be overparameterized or overfitted as they include many variables in order to provide a model that can be generalized over the whole data. This generalization approach would lead to overfitting with linear regression as the latter solves the weights by finding the inverse of the correlation matrix.

### Limitations

The main limitation of our analysis is that it is a post-hoc analysis of a trial with negative results on primary outcome^6^. Given the changes in practice over time, our findings should be considered as exploratory in nature and applicable to the setting of the trial^6^. Further, our findings might have been affected by sample size being smaller than the cardiac trials or trials of clinical endpoints^27^. In our initial sample size calculation, we had cautiously planned 80% power to detect 4.5% difference in future FRS^36^. Other investigators were more optimistic and planned for 20% difference in FRS^7^. It is likely that any future trial incorporating this methodology should be performed in rural areas or areas of need, at large distances from tertiary teaching hospitals and include a much smaller effect size than the 4.5% effect size that we used. Identification of areas of need can be performed using data driven approaches to assess geographical variation rather than a simple rural metropolitan divide ^37^.

## Conclusion

Future FRS is largely determined at trial entry. Given that the major determinants of future FRS are not modifiable, other approaches to secondary prevention trial such as using clinical endpoints should be explored. Our findings have implications for the use of FRS as the primary outcome in clinical trials targeting stroke prevention.

## Data Availability

The data is available on written request to Dr Amanda Thrift

## Funding

The STANDFIRM trial was supported by a National Health and Medical Research Council (NHMRC) project grant (586605) and fellowship support from the NHMRC for AGT (1042600) and DAC (1063761)

Supplementary Figure 6: SVR with baseline FRS removed. The predicted values are underestimated at higher range of the observed values. FRS, Framingham risk score; IHD, ischemic heart disease, AF, atrial fibrillation; HbA1c, glycosylated hemoglobin.

